# Distinguishing between COVID-19 and the common cold in a primary care setting - comparison of patients with positive and negative SARS-CoV-2 PCR results

**DOI:** 10.1101/2020.04.27.20081877

**Authors:** Johannes Just, Marie-Therese Puth, Felix Regenold, Klaus Weckbecker, Markus Bleckwenn

## Abstract

**Background:** Combating the COVID-19 pandemic is a major challenge for health systems, citizens and policy makers worldwide. Early detection of affected patients within the large and heterogeneous group of patients with common cold symptoms is an important element of this effort, but often hindered by limited testing resources and the lack of pathognomonic symptoms in COVID-19. Therefore, we aimed to identify predictive risk factors for a positive SARS-CoV-2 PCR (CovPCR) result in a primary care setting.

**Method:** We performed a multi-center cross-sectional cohort study on predictive clinical characteristics for a positive CovPCR over a period of 4 weeks in primary care patients in Germany.

**Findings:** In total, 374 patients in 14 primary care centers received CovPCR and were included in this analysis. The median age was 44.0 (IQR: 31.0-59.0) and a fraction of 10.7% (n=40) tested positive for COVID-19. Patients who reported anosmia had a higher odds ratio (OR: 4.54; 95%-CI: 1.51-13.67) for a positive test result while patients with a sore throat had a lower OR (OR: 0.33; 95%-CI: 0.11-0.97). Patients who had a first grade contact with an infected persons and showed symptoms themselves also had an increased OR for positive testing (OR: 5.16; 95% CI: 1.72-15.51). This correlation was also present when they themselves were still asymptomatic (OR: 12.55; 95% CI: 3.97-39.67).

**Conclusions:** Several anamnestic criteria may be helpful to assess pre-test probability of COVID-19 in patients with common cold symptoms

## Background

The COVID-19 pandemic is a major challenge for health systems, citizens and policy makers worldwide [1]. The early political implementation of individual distancing has slowed down the reproduction rate of the virus in Germany significantly [2]. In addition, compared to other countries, Germany has implemented an intensive testing strategy for new infections using PCR swab testing, funded by the national social health insurance system (statutory health insurance). According to date from “Robert-Koch Institut” (RKI), roughly an equivalent to the CDC in the US, 176 participating laboratories reported a number of 348,619 PCR tests carried out in calendar week 12/2020 (419 tests/100.000 inhabitants). The tests were mostly carried out in doctors’ (GP’s) practices, newly formed COVID-19 test centers and hospitals [3].

In view of the potentially exponential increase in the number of new cases and finite laboratory resources (test stations, reagents, etc.), there is an urgent need to collect and evaluate clinical data on the clinical features of tested patients. Of particular interest is the question of how far positive and negative patients differ in their initial clinical presentation. The more precisely physicians use SARS-CoV-2 PCR (CovPCR) testing, the more efficient it is. Statistically, this is reflected in Baye’s theorem: The quality of a test is not only determined by specificity and sensitivity, but also depends on the pre-test probability of the event to be tested. Thus, if the pre-test probability for COVID-19 is higher, the probability that a CovPCR-positive patient is actually ill (positive-predictive value = PPV) is increased [4].

In this paper we aimed to identify predictive risk profiles for a positive CovPCR result in primary care.

## Method

### Study design

In this multi-center, cross-sectional study, the characteristics of patients who tested positive and negative for COVID-19 were assessed. A total of 26 office-based specialists for internal and/or general medicine with a full primary care mandate from 14 different locations participated in the study.

### Setting

All locations collected patient-related data based on a uniform quality standard in the documentation of COVID-19 suspect cases that we provided. Each site reported anonymous data on all CovPCR swabs taken. All data were noted on a paper-based, structured documentation form and passed on anonymously for evaluation. The documentation form was prepared on the basis of the recommendations of the RKI as well as the available evidence on symptoms and risk factors for COVID-19 and was subjected to pre-testing with experienced family physicians [5-8]. The test reason “Patient is contact person without symptoms” was included in addition to the test reasons recommended by the RKI, as it had already been used frequently in everyday practice. The study period was from 24.03.2020 to 17.04.2020 - this period was chosen because the RKI criteria for carrying out a CovPCR remained unchanged.

### Participants

The participating GP’s practices represented a “convenience sample”. Selection criteria were the performance of smear tests, structured documentation and willingness to participate. All patients who received CovPCR in the participating GP’s practices within the study period were eligible for inclusion. Patients whose tests had been carried out for procedural reasons and did not correspond to a specific clinical indication were excluded (e.g. testing of recovered patients after end of quarantine). There were no other exclusion criteria.

### Variables and data sources

The variables under study included: Age, sex, reason for the test, risk factors, symptoms leading to the test, result expected by the GP and actual CovPCR results. All information was extracted from the anonymized documentation forms

### Bias

In order to ensure that there was no excessive distortion in the patient collective due to the convenience sampling of practices, age and sex of the tested persons were compared with demographic data from a large national sample of CovPCR-tests [3].

### Statistical methods

Data entry was performed twice and the data sets were compared digitally, any deviations were checked and a plausibility check of the data entered was performed. The statistical evaluation was carried out with the statistics program R. In addition to the descriptive statistics, initial chi-square tests were performed for different potential influencing factors (symptoms, test reason, etc.) on the test result (CovPCR positive yes/no). Statistically significant influencing factors were then transferred to a logistic regression model to check their effect strength. Missing data were reported in the descriptive part, for the regression model missing values were assigned to the more frequent result (deviation to the middle), a proportion of missing values >5% was not exceeded per variable.

The study design was reviewed by the ethics committee of the University of Leipzig, Germany under the procedure number 184/20-ek, there were no ethical concerns. This study was conducted without external financial support.

## Results

The participating practices tested n=374 patients per CovPCR in the investigated period. Of these, 10.7% (n=40) tested positive. The symptom anosmia, as well as contact with infected persons, was associated with a positive CovPCR result.

A total of 26 specialists with a general practitioner mandate at 14 different locations (individual and group practices) from North Rhine-Westphalia, Rhineland-Palatinate, Hesse and Saxony-Anhalt (four of 16 federal German states) took part in the study. All practices are located in urban and rural districts with moderate to increased COVID-19 activity (cumulative incidence 50-250 cases per 100,000 inhabitants) [2].

Parallel to the number of new infections reported throughout Germany, the number of tests performed decreased in the course of the study, as did the rate of positively tested patients.

**Figure 1:**
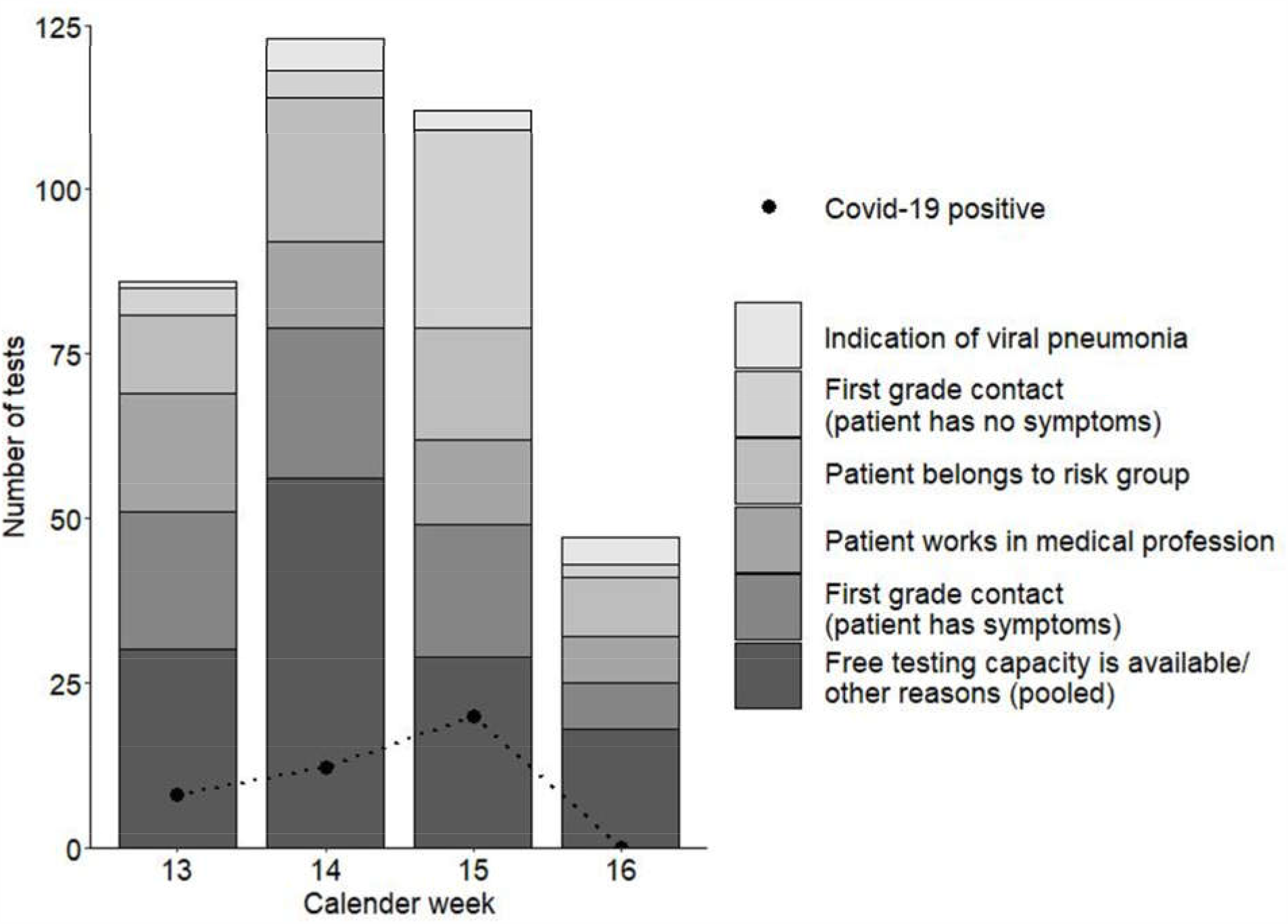
Number of tests separated by test reason per calendar week

Of the examined patients, 58.5% were female and the median age was 44 years. Further characteristics of the investigated cohort can be found in table 1.

**Table 1:**
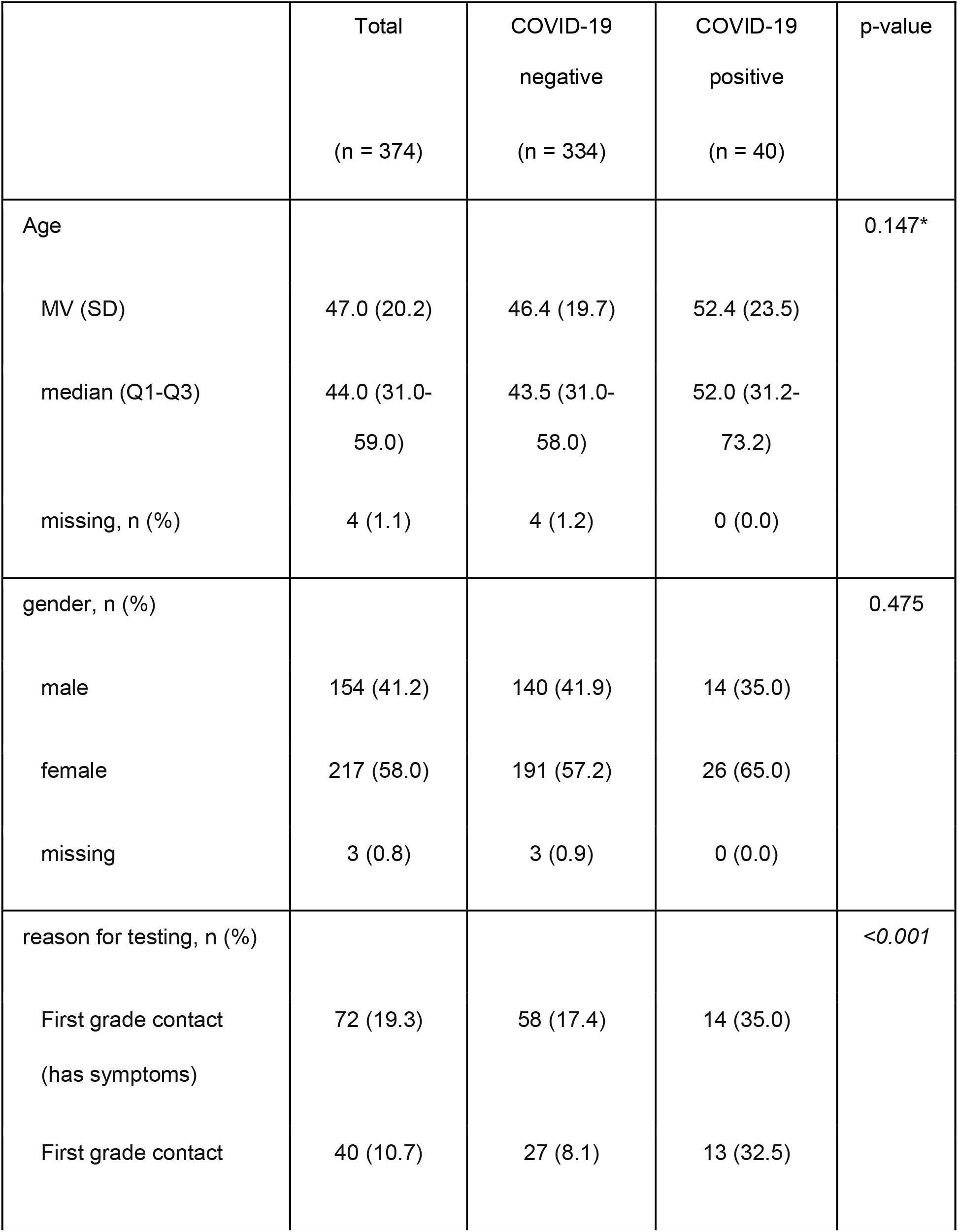
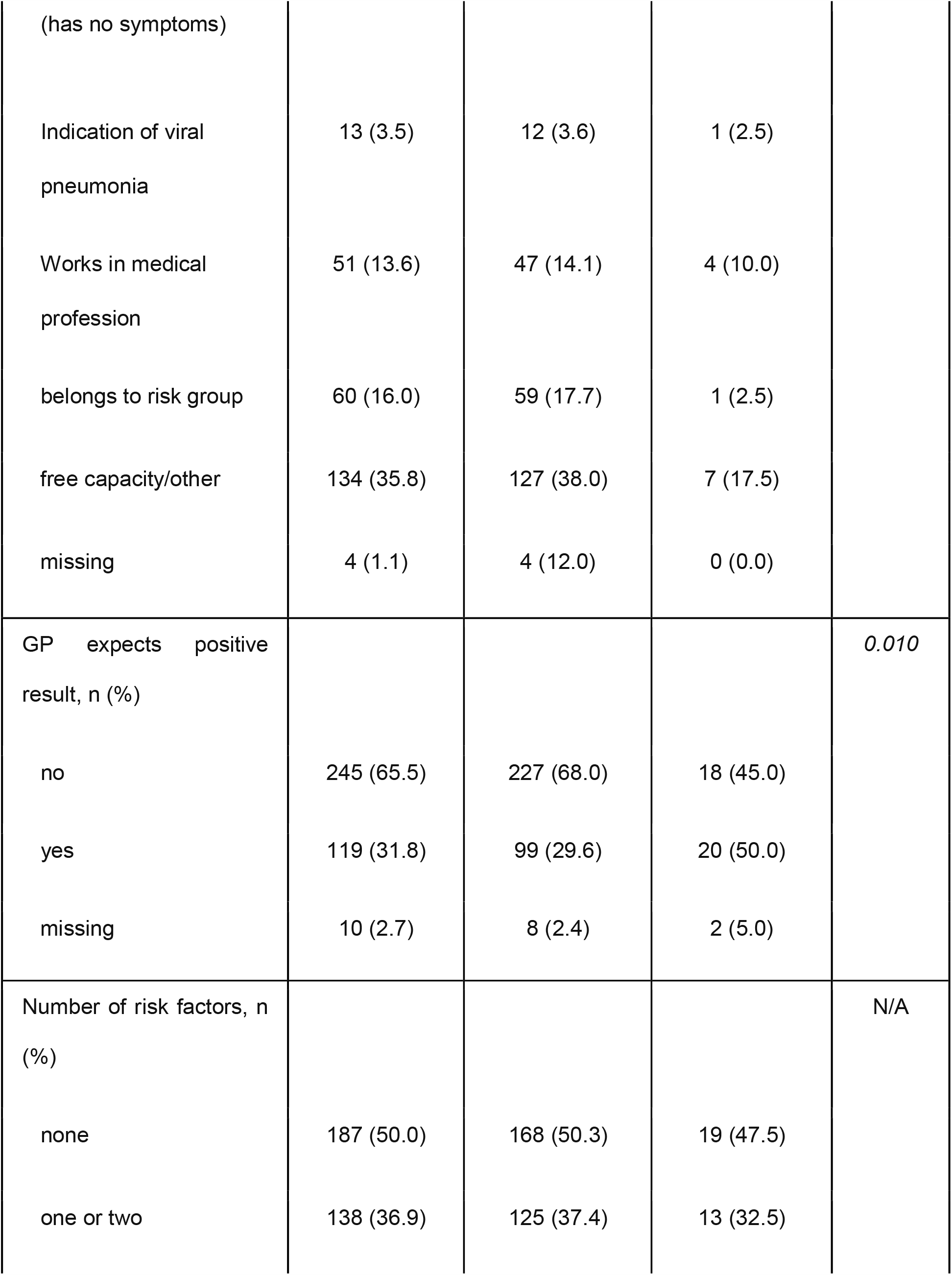
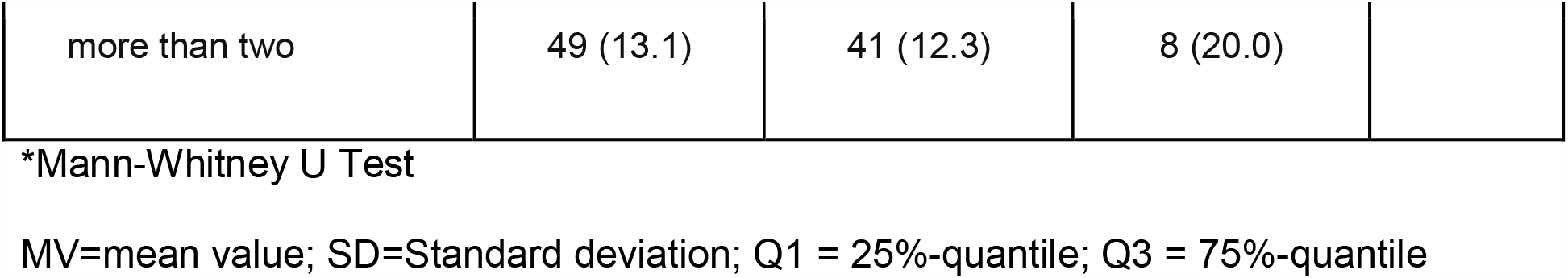
Patient characteristics

The most frequent symptoms at the presentation of patients who later tested positive for COVID-19 were cough, fever, anosmia and muscle pain. The most common symptoms at presentation of patients who later tested negative for COVID-19 were cough, sore throat, fatigue and fever. A complete list of symptoms is displayed in table 2.

**Table 2:** Symptoms of patients tested negative and positive for COVID-19

| Variable | COVID-19 negative<br>(n = 307) |  | COVID-19 positive<br>(n = 27) |  | p-value |
| --- | --- | --- | --- | --- | --- |
| cough (n, %) | 214 | 69.7 | 19 | 70.4 | 1.000 |
| Sore throat (n, %) | 120 | 39.1 | 5 | 18.5 | 0.038 |
| fatigue (n, %) | 89 | 29.0 | 5 | 18.5 | 0.371 |
| fever (n, %) | 84 | 27.4 | 9 | 33.3 | 0.507 |
| nasal congestion (n, %) | 84 | 27.4 | 5 | 18.5 | 0.373 |
| muscle pain (n, %) | 59 | 19.2 | 7 | 25.9 | 0.448 |
| dyspnea (n, %) | 56 | 18.2 | 4 | 14.8 | 0.798 |
| headache (n, %) | 47 | 15.3 | 3 | 11.1 | 0.780 |
| anorexia (n, %) | 28 | 9.1 | 2 | 7.4 | 1.000 |
| diarrhea (n, %) | 23 | 7.5 | 1 | 3.7 | 0.707 |
| anosmia (n, %) | 22 | 7.2 | 7 | 25.9 | 0.005 |
| chills (n, %) | 20 | 6.5 | 5 | 18.5 | 0.040 |
| nausea (n, %) | 11 | 3.6 | 0 | 0.0 | 1.000 |
| vomiting (n, %) | 4 | 1.3 | 0 | 0.0 | 1.000 |
| other symptoms<br>(n, %) | 37 | 12.1 | 3 | 11.1 | 1.000 |

Asymptomatic patients (n=40) were excluded in this table.

The regression model used to determine the individual effect sizes included all patient characteristics that showed a statistically significant influence on the test result in the Chi-square test. The odds ratio (OR) for a positive CovPCR was increased in patients who had contact with an infected person who were older and in patients with anosmia and decreased in patients suffering from a sore throat. Table 3 shows all results of the regression model.

**Table 3:** Logistic regression model: factors influencing positive CovPCR

| Variable | COVID-19<br>negative<br>(n=330)<br>n (%) | COVID-19<br>positive<br>(n=40)<br>n (%) | adjusted<br>OR<br>(95% CI) | p-value |
| --- | --- | --- | --- | --- |
| First grade contact<br>(has symptoms) | 58 (17.5) | 14 (35.0) | 5.16<br>(1.72-15.51) | 0.002 |
| First grade contact<br>(has no symptoms) | 27 (11.2) | 13 (32.5) | 12.55<br>(3.97-39.67) | <0.001 |
| Free capacity/other | 125 (37.9) | 7 (17.5) | 1.50<br>(0.46-4.87) | 0,497 |
| age* | 46,4 (19.7) | 52,4 (23.5) | 1.02<br>(1.00-1.04) | 0,019 |
| GP expects positive result | 97 (29.4) | 20 (50.0) | 1.98<br>(0.90-4.38) | 0.092 |
| chills | 20 (6.1) | 5 (12.5) | 2.80<br>(0.83-9.43) | 0.117 |
| Anosmia | 22 (6.7) | 7 (17.5) | 4.54<br>(1.51-13.67) | 0.011 |
| sore throat | 118 (35.8) | 5 (12.5) | 0.33<br>(0.11-0.97) | 0.029 |

OR = odds ratio; CI = confidence intervals
\* mean value (standard deviation); Nagelkerke R<sup>2</sup> 0.265

## Discussion

### Main result

The main result of this work is the shown correlation between a positive CovPCR and the symptom anosmia as well as a confirmed contact with an infected person, regardless of whether the tested person itself had symptoms or not.

### Strengths and limitations

In this work, predictive factors for a positive CovPCR were investigated for the first time. The evaluation was performed with a multicentric approach and was based on a common documentation standard. The selection of the practices was non-randomized, as a rapid implementation of the study was preferred to a lengthier randomized selection in respect to the pandemics’ rapid evolution. This may have resulted in a distortion of the investigated patient collective, although, no conspicuous deviations were found in comparison to the demographic data of all patients tested in Germany as published by the RKI: the age median in our population was 44 years (IQR: 31-59) compared to 42 years (IQR: 29-56) in a sample of +1.000.000 patients tested in Germany [3]. Therefore, the results could be considered representative for patients in Germany. The major limitation of this study is the small sample size resulting in wide CIs and a connected uncertainty regarding the strength and direction of several effects. Still, this is the first study to show clinical risk factors for a positive CovPCR and its results are in line with the current evidence base [5-8].

### Number of patients tested and rates of positives

The proportion of test-positive patients in our population was 10.7%, which is slightly above the average of positive tests performed in Germany during this period (North Rhine-Westphalia (9.3%), Rhineland-Palatinate (9.6%), Hesse (11.8%)) [3]. The number of tests decreased towards the end of the investigation period, which parallels the decreasing number of new COVID-19 infections in Germany as well as the end of the pandemic cold season in early spring.

### Factors that were associated with a negative test result

Negative tests were associated with the symptom “sore throat”. This result is consistent with the low prevalence of the symptom “sore throat” in confirmed COVID-19 cases [6]. The correlation found between age and test results is marginal (younger people were tested negative more often) and may be explained by local outbreaks in nursing homes. This effect will most likely disappear due to diffusion processes during the course of the pandemic and may not be present in countries with a more homogenous spreading pattern.

### Factors that were associated with a positive test result

Positive tests were associated with confirmed contact. This was to be expected, but it was surprising that this also applied to asymptomatic patients. He et al. concluded that 45% of infections occur in the pre-symptomatic phase of the patient, i.e. the patient is still asymptomatic but already has large amounts of virus replicated in the throat and is highly contagious [9]. This result underlines the particular relevance of contact anamnesis for the test decision, but also the relevance of early isolation of potentially affected contacts.

Of particular clinical importance is the observed association between the symptom anosmia and a positive PCR result. Anosmia has already been described as a relevant COVID-19 symptom in other studies, but this is the first study to show that anosmia may be a useful discriminator between COVID-19 and other, endemic respiratory tract infections [8,10,11]. Since the examined patients mainly presented with fever, cough, muscle pain and other cold symptoms, which are common with endemic respiratory tract infections, anosmia can be an important indication of COVID-19. Although anosmia is a secondary symptom of a variety of viral respiratory infections, the incidence of anosmia without acute or chronic rhinitis in COVID-19 is striking [8,10-12]. Reporting from our clinical experience with COVID-19, anosmia was sometimes so pronounced in COVID-19 patients that it was presented as the sole symptom upon presentation. Therefore, we recommend that patients are always asked for fever, cough, dyspnea and anosmia as part of the initial stratification process. In case of isolated anosmia in the absence of chronic or acute rhinosinusitis, we recommend a testing for COVID-19 infection, as already suggested in a French case series by Villalba et al. and others [8,11,13]

## Conclusion

Rapid testing of first degree contacts, consistent tracing of infection chains and quarantine will remain essential parts of the COVID-19 containment strategy. Our data show that the testing of all first-degree contacts, including asymptomatic ones can be of merit and that anosmia, even as an isolated symptom is a clear indication of COVID-19 infection.

## Data Availability

Data is available from the authors upon request within the framework of the basic data protection regulation

## List of abbreviations

CovPCR: SARS-CoV-2 PCR
PPV: positive-predictive value

## Declarations

Ethics approval and consent to participate:

The study design was reviewed by the ethics committee of the University of Leipzig, Germany under the procedure number 184/20-ek, there were no ethical concerns. Consent to participate was not applicable according to national regulations as routine treatment data was anonymously passed on to researchers. The ethics committee stated no concerns regarding the method of data collection including consent to participate.

### Consent for publication

All authors gave their consent for publication

### Availability of data and materials

The datasets generated and analysed during the current study are available from the corresponding author on reasonable request.

### Authors contributions

JJ, KW and MB designed the study, supervised data collection and drafted main parts of the paper. FR gathered data, performed statistical analysis and contributed to the development of the paper. MP performed statistical analysis and helped drafting the paper. All authors read and approved the final manuscript.

## Acknowledgements

We thank all participating practices for their effort.

